# Amyloid-β PET and CSF in an autopsy confirmed cohort

**DOI:** 10.1101/2020.06.02.20119891

**Authors:** Juhan Reimand, Baayla D.C. Boon, Lyduine E. Collij, Charlotte E. Teunissen, Annemieke J.M. Rozemuller, Bart N.M. van Berckel, Philip Scheltens, Rik Ossenkoppele, Femke Bouwman

**Affiliations:** Department of Neurology & Alzheimer Center Amsterdam, Amsterdam Neuroscience, Vrije Universiteit Amsterdam, Amsterdam UMC, Amsterdam, the Netherlands.; Department of Health Technologies, Tallinn University of Technology, Tallinn, Estonia; Radiology Centre, North Estonia Medical Centre, Tallinn, Estonia; Department of Pathology, Amsterdam Neuroscience, Vrije Universiteit Amsterdam, Amsterdam UMC, Amsterdam, the Netherlands.; Department of Radiology and Nuclear Medicine, Amsterdam Neuroscience, Vrije Universiteit Amsterdam, Amsterdam UMC, Amsterdam, the Netherlands.; Neurochemistry Laboratory, Department of Clinical Chemistry, Amsterdam Neuroscience, Vrije Universiteit Amsterdam, Amsterdam UMC, Amsterdam, the Netherlands; Clinical Memory Research Unit, Lund University, Lund, Sweden

## Abstract

**Objective:** Accumulation of amyloid-β is among the earliest changes in Alzheimer’s disease (AD). Amyloid-β positron emission tomography (PET) and Aβ_42_ in cerebrospinal fluid (CSF) both assess amyloid-β pathology *in-vivo*, but 10–20% of cases show discordant (CSF+/PET- or CSF-/PET+) results. The neuropathological correspondence with amyloid-β CSF/PET discordance is unknown.

**Methods:** We included 21 patients from our tertiary memory clinic who had undergone both CSF Aβ_42_ analysis and amyloid-β PET, and had neuropathological data available. Amyloid-β PET and CSF results were compared with neuropathological ABC scores (comprising of Thal (A), Braak (B) and CERAD (C) stage, all ranging from 0 [low] to 3 [high]) and neuropathological diagnosis.

**Results:** Neuropathological diagnosis was AD in 11 (52%) patients. Amyloid-β PET was positive in all A3, C2 and C3 cases and in one of the two A2 cases. CSF Aβ_42_ was positive in 92% of A2/A3 and 90% of C2/C3 cases. PET and CSF were discordant in 3/21 (14%) cases: CSF+/PET- in a patient with granulomatosis with polyangiitis (A0B0C0), CSF+/PET- in a patient with FTLD-TDP type B (A2B1C1), and CSF-/PET+ in a patient with AD (A3B3C3). Two CSF+/PET+ cases had a non-AD neuropathological diagnosis, i.e. FTLD-TDP type E (A3B1C1) and adult-onset leukoencephalopathy with axonal spheroids (A1B1C0).

**Interpretation:** Our findings confirm amyloid-β CSF/PET discordance with a range of possible reasons. Furthermore, amyloid-β biomarker positivity on both PET and CSF did not invariably result in an AD diagnosis at autopsy, illustrating the importance of considering relevant co-morbidities when evaluating amyloid-β biomarker results.

## Introduction

Among the earliest neuropathological events in Alzheimer’s disease (AD) is the accumulation and aggregation of amyloid-β, which occurs decades before symptom onset.^1^ Amyloid-β can aggregate in the brain parenchyma as plaques or in the cerebral vasculature as cerebral amyloid angiopathy (CAA). Two methods are currently employed to assess amyloid-β pathology *invivo*. Aβ_42_ levels in the cerebrospinal fluid (CSF) reflect the concentrations of soluble amyloid-β, which has been shown to correlate with amyloid-β deposits in the brain.^2^ Alternatively, positron emission tomography (PET) with amyloid-β radiotracers can be used to visualize cerebral amyloid-β deposits.^3–5^ These two methods are considered interchangeable for the assessment of amyloid pathology *in-vivo* and for the diagnosis of AD in both clinical practice and research.^6,7^

In the majority of cases amyloid-β PET and CSF are concordant, but in 10–20% of patients they show discordant (CSF+/PET- or CSF-/PET+) results.^8,9^ One possible hypothesis for the amyloid-β CSF/PET discordance is that soluble CSF Aβ_42_ decreases before significant fibrillar amyloid-β deposits can be detected by PET.^10,11^ Although studies have been performed to compare either amyloid-β PET or CSF Aβ_42_ to neuropathological examination results,^2–5^ so far no head-to-head cohort studies have been performed to compare *in-vivo* amyloid-β CSF and PET results to neuropathological findings. Previously, two case reports of patients with discordant amyloid-β CSF/PET (both CSF+/PET-) and available neuropathology have been published,^12,13^ in which the negative PET signal was attributed to the lack of neuritic plaques at autopsy. Further investigating the correspondence between amyloid-β PET, CSF and neuropathology (“standard of truth”) is important to shed light on the underlying cause of amyloid-β CSF/PET discordance. Also, if CSF+/PET- amyloid-β status is an indicator of early amyloid-β pathology, this could be instrumental for future disease modifying therapies.^14^ Therefore, the aim of this study was to investigate the concordance between PET and CSF amyloid-β status in a sample with available neuropathological results and to characterize the amyloid-β CSF/PET discordant cases neuropathologically.

## Materials and methods

### Participants

We retrospectively included 21 autopsy cases from the Amsterdam Dementia Cohort who had undergone both CSF Aβ_42_ analysis and amyloid-β PET during life. Patients visiting our tertiary memory center are screened according to a standardized protocol,^15^ including a clinical and neuropsychological evaluation, *APOE* genotyping, magnetic resonance imaging (MRI), and a lumbar puncture (LP) for CSF biomarker analysis. Clinical diagnosis is determined during a multidisciplinary meeting.

Amyloid-β PET and CSF analyses were performed between 2007 and 2016 and neuropathological diagnosis was performed between 2011 and 2019 (Fig 1). In this sample, LP for CSF analysis always preceded amyloid-β PET and, the median CSF-PET time was 28 [interquartile range (IQR): 18, 56] days. The median time difference between amyloid-β PET and patient death was 3.0 (IQR: 1.7, 6.5) years and the time difference between LP and patient death was 3.3 (IQR: 2.0, 6.7) years.

**Figure 1.**
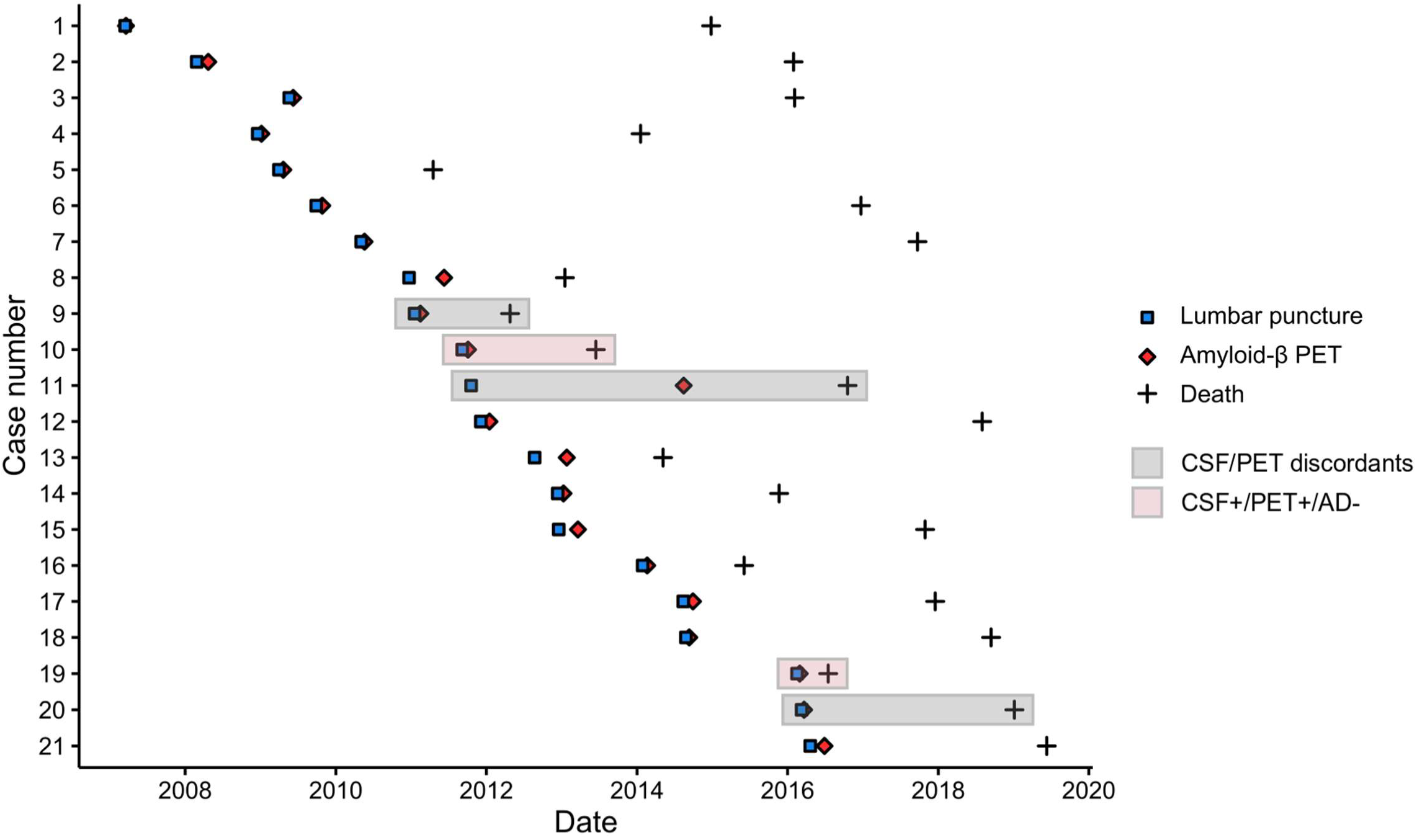
Time between lumbar puncture, amyloid-β PET and patient death. Abbreviations: AD Alzheimer’s disease, CSF cerebrospinal fluid, PET positron emission tomography.

### Standard protocol approvals and patient consents

The institutional review board of the VU University Medical Center approved all studies from which the current data was gathered and retrospectively analyzed. All patients provided written informed consent for their data to be used for research purposes.

### Cerebrospinal fluid

CSF was obtained during life by LP between L3/4 and L5/S1, using a 25-gauge needle and a syringe.^16^ Samples were collected in polypropylene collection tubes and centrifuged at 1800g for 10 min at 4°C and thereafter frozen at –20 °C until routine biomarker analysis. Manual analyses of Aβ_42_, total tau (t-tau) and phosphorylated tau (p-tau) were performed using sandwich ELISAs (Innotest assays: β-amyloid1-42, tTAU-Ag and PhosphoTAU-181p; Fujirebio) in the Neurochemistry Laboratory of the Department of Clinical Chemistry of Amsterdam UMC. If two CSF results were available, we used the result closest to the amyloid-β PET (3 cases, all with concordant Aβ_42_ status between two samples). As median CSF Aβ_42_ values of our cohort have been gradually increasing over the years, we were unable to use the original CSF Aβ_42_ values.^17^ Therefore, we used CSF Aβ_42_ values that have been adjusted for the longitudinal upward drift with a uniform cut-off of 813 pg/ml (< 813 pg/mL considered as CSF amyloid-β positive).^18^ Additionally, as it has been previously shown that the ratio of CSF Aβ_42_ with CSF (p)tau is superior to CSF Aβ_42_ in predicting the diagnosis of AD,^19^ we also used a CSF p-tau/Aβ_42_ ratio with a previously validated cut-off of 0.054.^20^ This cut-off was obtained by mixture modelling of 2711 CSF results of the Amsterdam Dementia Cohort, similar to previous work.^18^

### Amyloid-β positron emission tomography

Amyloid-β PET was performed using the following PET scanners: ECAT EXACT HR+ scanner (Siemens Healthcare, Germany), Gemini TF PET/CT, Ingenuity TF PET-CT and Ingenuity PET/MRI (Philips Medical Systems, the Netherlands). We included fifteen cases with [^11^C]Pittsburgh Compound-B (PIB),^21^ three with [^18^F]florbetaben^22^ and three with [^18^F]flutemetamol.^23^ Amyloid-β PET status (positive or negative) was determined by a majority visual read of three reads. All scans were initially read by an expert nuclear medicine physician (BvB, from 2007 to 2016, read 1). In addition, in 2019 the scans were reread for this study by BvB (read 2) and LC (with extensive experience in reading amyloid-β PET scans, read 3), while being blinded to the results of other visual reads, CSF, and neuropathological results. The three amyloid-β PET visual reads were concordant (either +/+/+ or -/-/-) in 18/21 cases. In the three remaining cases (nr 5, 10, 19), two of the three visual reads were positive, and as such these cases were considered PET-positive.

### Neuropathology

Autopsies were performed by the Department of Pathology of Amsterdam UMC; location VUmc for the Netherlands Brain Bank or for VUmc. Brain donors or their next of kin signed informed consent regarding the usage of brain tissue and clinical records for research purposes. Brain autopsies and neuropathological diagnosis were performed according to international guidelines of Brain Net Europe II consortium (http://www.brainnet-europe.org) and the applicable diagnostic criteria.^24,25^ For this particular study, every case also without suspicion of AD pathology during life, was scored by AR and BB for AD neuropathological changes according to the ABC scoring system by AR and BB,^24^ in which the A stands for amyloid-β Thal phase,^26^ B for Braak stage for neurofibrillary tangles,^1^ and C for CERAD criteria for neuritic plaques.^27^ When present, CAA was classified as Type 1 (including capillaries in the parenchyma) or Type 2 (leptomeningeal/cortical without capillary involvement) and staged according to Thal et al.^28^

### Statistical analysis

Statistical analysis was conducted using R software (Version 3.6.1). We used descriptive statistics to characterize the sample. We used the Cochrane-Armitage trend test to examine the associations between amyloid-β biomarkers and the neuropathological ABC scores,^29^ which allowed us to compare both PET and CSF to neuropathology as we had only binarized results available for amyloid-β PET.

## Results

### Study population

In our sample of 21 cases, 16 (76%) were male and 10 (48%) were carriers of an *APOE* ε4 allele (Table 1). Mean age at death was 65±8 years and the average last known Mini-Mental State Examination (MMSE, median 2.0 years before death) was 20±6. Eleven (52%) patients had a clinical diagnosis of AD, which was in accordance with neuropathological diagnosis in all AD cases. In 15 (71%) cases CAA (11 CAA-Type 1, 4 CAA-Type 2) was observed at neuropathological examination.

**Table 1.**
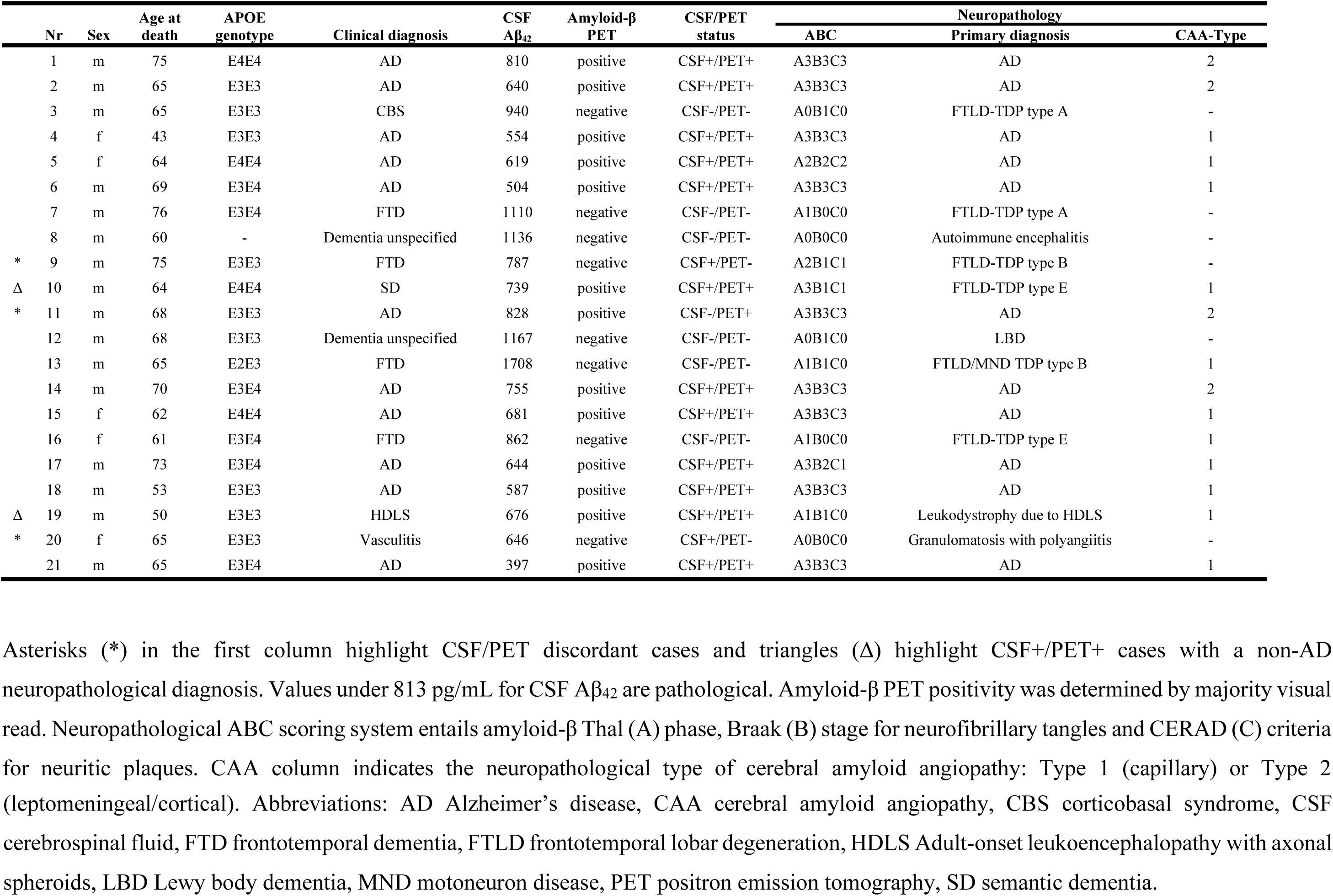
Case characteristics.

| Nr | Sex | Age at death | APOE genotype | Clinical diagnosis | CSF Aβ<sub>42</sub> | Amyloid-β PET | CSF/PET status | Neuropathology | | |
| --- | --- | --- | --- | --- | --- | --- | --- | --- | --- | --- |
|  |  |  |  |  |  |  |  | ABC | Primary diagnosis | CAA-Type |
| 1 | m | 75 | E4E4 | AD | 810 | positive | CSF+/PET+ | A3B3C3 | AD | 2 |
| 2 | m | 65 | E3E3 | AD | 640 | positive | CSF+/PET+ | A3B3C3 | AD | 2 |
| 3 | m | 65 | E3E3 | CBS | 940 | negative | CSF-/PET- | A0B1C0 | FTLD-TDP type A | - |
| 4 | f | 43 | E3E3 | AD | 554 | positive | CSF+/PET+ | A3B3C3 | AD | 1 |
| 5 | f | 64 | E4E4 | AD | 619 | positive | CSF+/PET+ | A2B2C2 | AD | 1 |
| 6 | m | 69 | E3E4 | AD | 504 | positive | CSF+/PET+ | A3B3C3 | AD | 1 |
| 7 | m | 76 | E3E4 | FTD | 1110 | negative | CSF-/PET- | A1B0C0 | FTLD-TDP type A | - |
| 8 | m | 60 | - | Dementia unspecified | 1136 | negative | CSF-/PET- | A0B0C0 | Autoimmune encephalitis | - |
| * 9 | m | 75 | E3E3 | FTD | 787 | negative | CSF+/PET- | A2B1C1 | FTLD-TDP type B | - |
| $\Delta$ 10 | m | 64 | E4E4 | SD | 739 | positive | CSF+/PET+ | A3B1C1 | FTLD-TDP type E | 1 |
| * 11 | m | 68 | E3E3 | AD | 828 | positive | CSF-/PET+ | A3B3C3 | AD | 2 |
| 12 | m | 68 | E3E3 | Dementia unspecified | 1167 | negative | CSF-/PET- | A0B1C0 | LBD | - |
| 13 | m | 65 | E2E3 | FTD | 1708 | negative | CSF-/PET- | A1B1C0 | FTLD/MND TDP type B | 1 |
| 14 | m | 70 | E3E4 | AD | 755 | positive | CSF+/PET+ | A3B3C3 | AD | 2 |
| 15 | f | 62 | E4E4 | AD | 681 | positive | CSF+/PET+ | A3B3C3 | AD | 1 |
| 16 | f | 61 | E3E4 | FTD | 862 | negative | CSF-/PET- | A1B0C0 | FTLD-TDP type E | 1 |
| 17 | m | 73 | E3E4 | AD | 644 | positive | CSF+/PET+ | A3B2C1 | AD | 1 |
| 18 | m | 53 | E3E3 | AD | 587 | positive | CSF+/PET+ | A3B3C3 | AD | 1 |
| $\Delta$ 19 | m | 50 | E3E3 | HDLS | 676 | positive | CSF+/PET+ | A1B1C0 | Leukodystrophy due to HDLS | 1 |
| * 20 | f | 65 | E3E3 | Vasculitis | 646 | negative | CSF+/PET- | A0B0C0 | Granulomatosis with polyangiitis | - |
| 21 | m | 65 | E3E4 | AD | 397 | positive | CSF+/PET+ | A3B3C3 | AD | 1 |

Asterisks (\*) in the first column highlight CSF/PET discordant cases and triangles ( $\Delta$ ) highlight CSF+/PET+ cases with a non-AD neuropathological diagnosis. Values under 813 pg/mL for CSF Aβ <sub>42</sub> are pathological. Amyloid-β PET positivity was determined by majority visual read. Neuropathological ABC scoring system entails amyloid-β Thal (A) phase, Braak (B) stage for neurofibrillary tangles and CERAD (C) criteria for neuritic plaques. CAA column indicates the neuropathological type of cerebral amyloid angiopathy: Type 1 (capillary) or Type 2
(leptomeningeal/cortical). Abbreviations: AD Alzheimer's disease, CAA cerebral amyloid angiopathy, CBS corticobasal syndrome, CSF cerebrospinal fluid, FTD frontotemporal dementia, FTLF frontotemporal lobar degeneration, HDLS Adult-onset leukoencephalopathy with axonal spheroids, LBD Lewy body dementia, MND motoneuron disease, PET positron emission tomography, SD semantic dementia.

### In vivo amyloid-β status

Thirteen (62%) cases were defined as amyloid-β positive based on PET, 14 (67%) based on CSF Aβ_42_ and 11 (52%) based on CSF p-tau/Aβ_42_ ratio. In our sample, CSF Aβ_42_ and amyloid-β PET were concordant in 18 (86%) cases. CSF p-tau/Aβ_42_ ratio was concordant with amyloid-β PET in 17 (81%) cases and with CSF Aβ_42_ in 16 (76%) cases.

### Discordance between amyloid-β PET, CSF and neuropathological diagnosis

Of the three discordant cases between CSF Aβ_42_ and amyloid-β PET, two were CSF+/PET- (case 9, clinical diagnosis: frontotemporal dementia [neuropathological diagnosis: frontotemporal lobar degeneration (FTLD)-TDP type B, ABC score: A2B1C1] and case 20 with vasculitis [granulomatosis with polyangiitis, A0B0C0], and one was CSF-/PET+ (case 11 with AD [AD, A3B3C3], Fig 2A-C). In addition, there were two CSF+/PET+ cases with a non-AD primary neuropathological diagnosis, i.e. case 10 with semantic dementia [FTLD-TDP type E, A3B1C1; CAA-Type 1 stage 2]; and case 20 with adult-onset leukoencephalopathy with axonal spheroids (HDLS) [leukodystrophy due to HDLS, A1B1C0; CAA-Type 1 stage 1] (Fig 2D-E).

**Figure 2.**
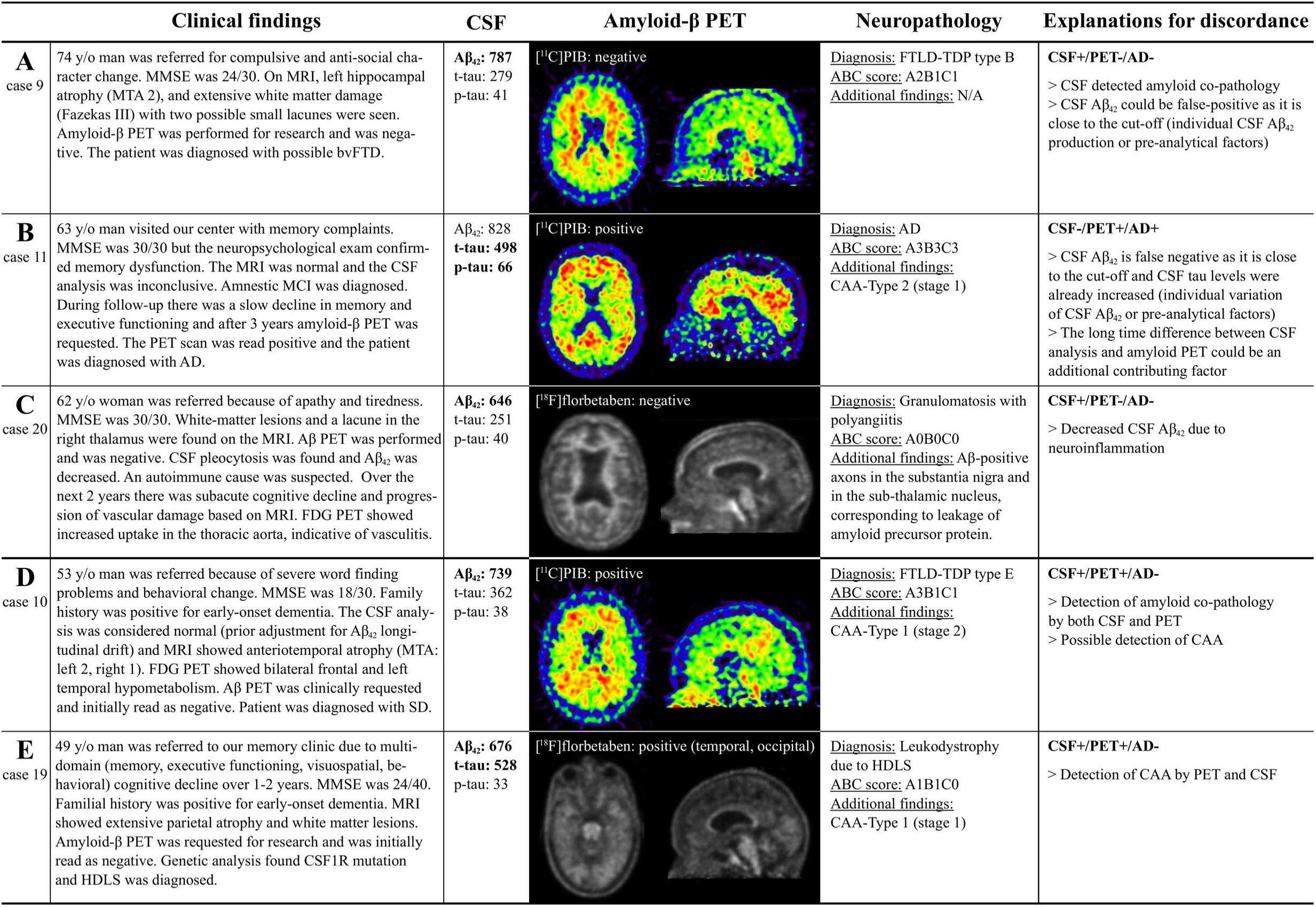
Discordance between amyloid-β CSF, PET and autopsy. Vignettes illustrating amyloid-β CSF/PET discordant cases (A,B,C) and CSF+/PET+ cases with a non-AD neuropathological diagnosis (D,E). CSF values for Aβ_42_ < 813 pg/mL, for phosphorylated tau (p-tau) > 52 pg/mL, and for total tau (t-tau) > 375 pg/mL are pathological (indicated by bold). Amyloid-β PET scans in cases 10 and 19 were initially read as amyloid-negative, but for this study the scans were considered amyloid-positive based on majority visual read. Abbreviations: AD Alzheimer’s disease, CAA cerebral amyloid angiopathy, CSF cerebrospinal fluid, FDG Fluorodeoxyglucose, FTD frontotemporal dementia, FTLD frontotemporal lobar degeneration, HDLS Adult-onset leukoencephalopathy with axonal spheroids, MCI Mild cognitive impairment, MMSE Mini-Mental State, Examination, MRI Magnetic resonance imaging, MTA – Medial temporal lobe atrophy, PET positron emission tomography, SD sematic dementia, TDP transactive response DNA binding protein.

### Association between biomarkers and ABC scores

CSF Aβ_42_ (Fig 3A) was positive in 12/13 (92%) and CSF p-tau/Aβ_42_ ratio (Fig 3B) in 10/13 (77%) of the A2/A3 cases. Both CSF Aβ_42_ and p-tau/Aβ_42_ ratio were positive in 10/11 (91%) B2/B3 cases and 9/10 (90%) C2/C3 cases. Amyloid-β PET (Fig 3C) was positive in one of the two A2 cases, and in all A3 and/or B2/B3 and/or C2/C3 cases. Cochrane trend analyses showed that there is an increasing proportion of biomarker-positive cases from score 0 to 3 across all ABC scores for amyloid-β PET (Z-score = –3.93 for A, Z = –3.81 for B, Z = –3.68 for C, all p < 0.001) and CSF Aβ_42_ (Z = –2.92, p = 0.003 for A; Z = –2.46, p = 0.014 for B; Z = –2.60, p = 0.009 for C).

**Figure 3.**
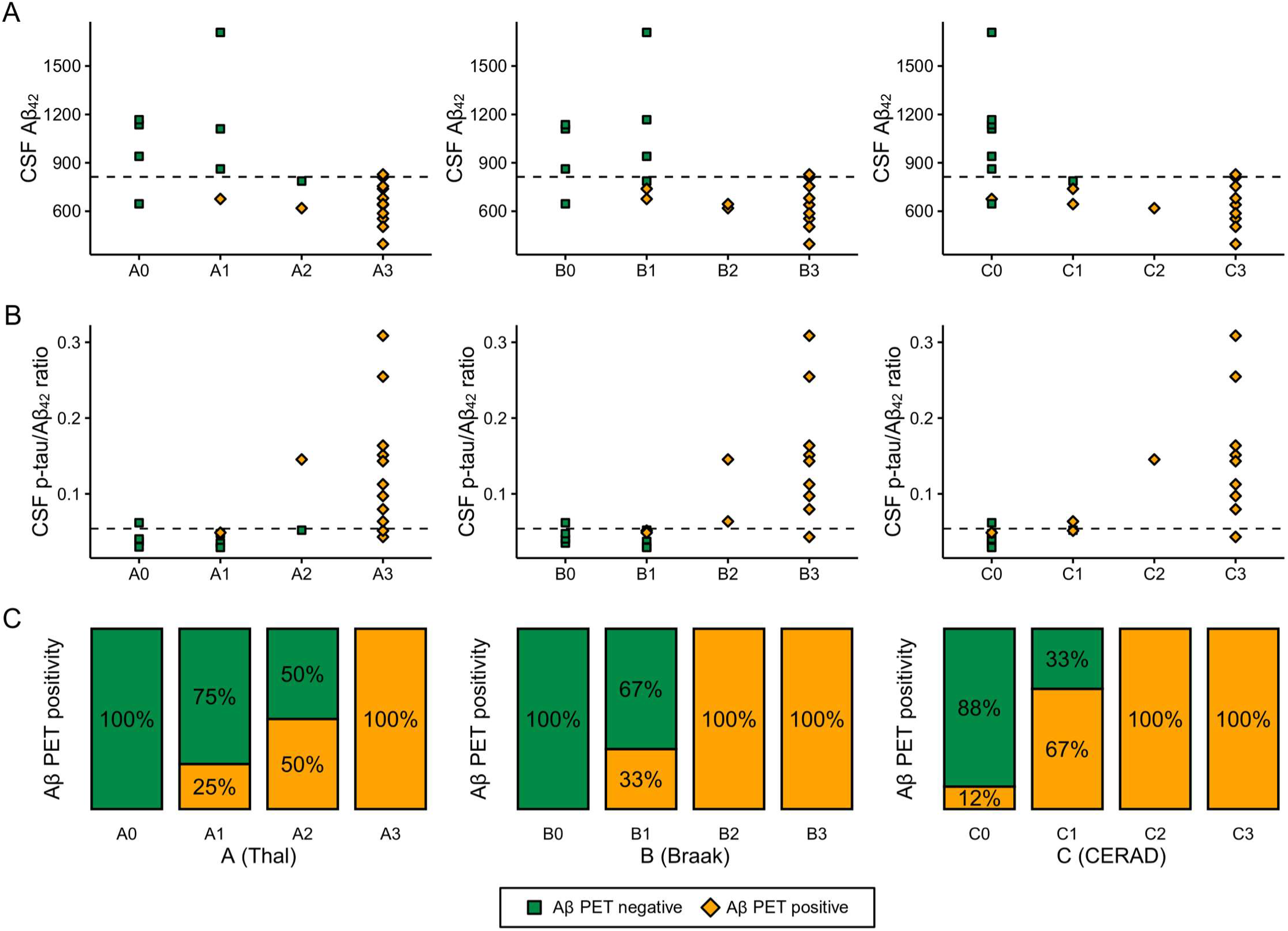
Correspondence of CSF Aβ_42_ (A), CSF p-tau/Aβ_42_ ratio (B), and amyloid-β (Aβ) PET (C) to neuropathological ABC scoring. Neuropathological ABC scoring system entails amyloid-β Thal phase (A0-A3), Braak stage for neurofibrillary tangles (B0-B3) and CERAD criteria for neuritic plaques (C0-C3). Dashed lines represent cut-offs for CSF Aβ_42_ (813 pg/mL) and CSF p-tau/Aβ_42_ ratio (0.054).

### Association between biomarkers and neuropathological diagnosis

Finally, we investigated the association between binarized biomarker results and neuropathological diagnosis. Amyloid-β PET was positive in all AD cases, but also indicated amyloid-β pathology in two cases without AD as neuropathological diagnosis (Fig 4). Both CSF Aβ_42_ and p-tau/Aβ_42_ were positive in 10/11 AD cases. Decreased CSF Aβ_42_ with a normal CSF p-tau/Aβ_42_ ratio was seen in three non-AD cases (HDLS [A1B1C0]. FTLD-TDP type B [A2B1C1], FTLD-TDP type E [A3B1C1]) and one AD case (A3B3C3). There were three cases with a non-AD neuropathological diagnosis (HDLS, FTLD-TDP type E, FTLD/MND-TDP type B) with normal levels of CSF p-tau but with increased CSF t-tau.

**Figure 4.**
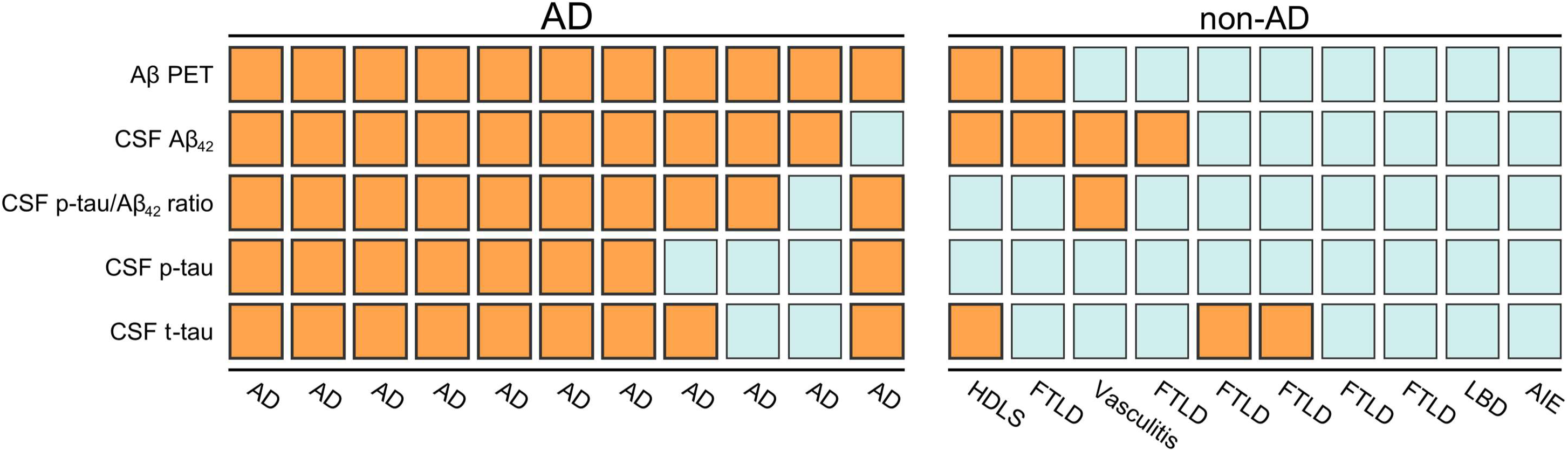
Biomarker status by primary neuropathological diagnosis. Colors indicate binarized status of biomarkers: orange for biomarker-positive, blue for biomarker-negative. Abbreviations: Aβ Amyloid-β, AD Alzheimer’s disease, AIE autoimmune encephalitis, CSF cerebrospinal fluid, FTLD frontotemporal lobar degeneration, HDLS Adult-onset leukoencephalopathy with axonal spheroids, PET positron emission tomography.

## Discussion

The primary aim of this study was to investigate the concordance between PET and CSF amyloid-β status in a sample with available neuropathological results in order to enhance our understanding of the amyloid-β CSF/PET discordant cases. We found that although both CSF and PET generally captured AD pathological change, there was still 14% (3/21) discordance between the two modalities. In our sample, possible reasons for amyloid-β CSF/PET discordance included neuroinflammation (CSF+/PET- in a case of granulomatosis with polyangiitis, A0B0C0), detection of amyloid-β co-pathology (CSF+/PET- in FTLD-TDP type B, A2B1C1) and additional factors influencing CSF Aβ_42_ levels (CSF-/PET+ in AD, A3B3C3). Additionally, we described two CSF+/PET+ non-AD cases illustrating that amyloid-β biomarker positivity on both PET and CSF does not invariably result in an AD diagnosis at autopsy. This highlights that it is important to consider other comorbidities when evaluating the results of amyloid-β biomarkers, especially since molecular biomarkers for non-AD neurodegenerative diseases are currently lacking.

Although in the majority of cases, amyloid-β PET and CSF Aβ_42_ show concordant results, 10–20% discordant CSF/PET status has repeatedly been shown.^8,9,30^ As amyloid-β CSF/PET discordance rates are highest in patients with early disease, it has been hypothesized that CSF/PET discordance might be partly explained by early decreases of CSF Aβ_42_ that precede amyloid-β depositions visible by PET.^10,11^ On the other hand, amyloid-β CSF/PET discordance in patients with dementia could be explained by one modality detecting beginning amyloid-β co-pathology in non-AD cases.^8^ To our knowledge, this is the first serial study including patients who have both amyloid-β PET and CSF Aβ_42_ in addition to neuropathological data available. In line with previous *in vivo* studies, we found a 14% (3/21) CSF/PET discordance rate. We reported a CSF+/PET- patient with A2B1C1 FTLD-TDP type B, where it is feasible that the reduction of CSF Aβ_42_ is caused by concomitant amyloid-β pathology. However, as the Aβ_42_ value was relatively close to the cut-off, it is not possible to entirely exclude individual CSF Aβ_42_ dynamics (i.e. this patient intrinsically producing less Aβ_42_)^31^ or pre-analytical factors.^32^ Previously, two CSF+/PET- case reports with available neuropathology have been published. First, a negative PIB PET scan was reported in a 91-year-old patient with abnormal CSF Aβ_42_ and tau biomarkers with sporadic AD.^12^ The negative amyloid-β PET status was attributed to the absence of a significant amount of fibrillar plaques (i.e. with a fibrillar core that the tracer binds to), although diffuse plaques were present. Second, low PIB PET retention with decreased CSF Aβ_42_ was reported in a familial AD case with arctic amyloid precursor protein (APP) mutation, thought to be caused by the lack of fibrillar amyloid-β plaques characteristic for this mutation.^13^ Future studies with neuropathological data are needed to further validate whether amyloid-β CSF+/PET- status is caused by beginning amyloid-β depositions and explore additional neuropathological substrates for CSF/PET discordance, such as differences in distribution, load and morphology of amyloid-β plaques and possible influences of co-pathologies.

In our sample, there were two cases with amyloid-β CSF+/PET+ biomarker status who did not meet neuropathological criteria for AD. The first had a diagnosis of FTLD-TDP type E with a high Thal score but only sparse neuritic plaques (A3B1C1). It is feasible that in this case both biomarkers detected concomitant amyloid-β co-pathology as increased PIB PET signal has been shown to be related to fibrillar plaque load even in case of sparse neuritic plaques.^33,34^ The patient was also diagnosed with CAA-Type 1 stage 2, which could also contribute to the amyloid-positivity, as CAA has been shown to affect both amyloid-β PET tracer uptake^35^ and CSF Aβ_42_ levels.^36^ The second CSF+/PET+ patient with a low score for AD pathology (A1B1C0) was diagnosed with HDLS, an autosomal dominant white matter disease due to mutations in the gene encoding colony stimulating factor 1 receptor (CSF1R).^37^ Previous case reports of HDLS including CSF analyses have provided no evidence for alterations in Aβ_42_ levels.^38,39^ It is also unlikely that pre-analytical assay effects caused the decrease of CSF Aβ_42_ in this case as the patient had a separate CSF Aβ_42_ sample with decreased Aβ_42_ four months earlier. Similar to the previous patient, CAA-Type 1 was present and might have contributed to the positive amyloid-β biomarker status, especially since PET tracer uptake was seen predominantly in the occipital region, a predilection site for CAA pathology.^35^ This illustrates that even concordant positivity of two amyloid-β biomarkers does not always result in a neuropathological diagnosis of AD, and relevant co-pathologies should always be considered.

There were four cases where CSF Aβ_42_ was decreased without a neuropathological diagnosis of AD. In three of them there was neuropathological evidence for amyloid-β co-pathology, but we also we described a CSF+/PET- case with granulomatosis with polyangiitis (formerly known as Wegener’s granulomatosis), which is in line with literature, as neuroinflammation^40,41^ as well as infection^42,43^ have been previously shown to cause decreased CSF Aβ_42_ without presence of AD pathology. This highlights that in select cases, there might be unspecific decreases in CSF Aβ_42_ levels without AD, although these cases might be distinguished from AD pathology based on clinical findings and MR imaging. In our particular case, after Aβ immunostaining, Aβ immunoreactive axons were seen, which can be attributed to the leakage of APP that is reported in various conditions such as ischemia, traumatic brain injury and – similar to this case – inflammation.^44^ The possible connection of this finding with the decrease of CSF Aβ_42_ is unclear, although it is tempting to hypothesize that the loss of APP leads to the interruption of the APP pathway and the reduction of its product Aβ_42_ in the CSF. We were unable to find previous case reports of vasculitis with available CSF Aβ_42_ analysis, but primary angiitis of the central nervous system has been associated with decreased APP in the CSF,^45^ lending support to that speculative theory.

CSF p-tau/Aβ_42_ ratio was slightly more specific than CSF Aβ_42_ for capturing the neuropathological diagnosis of AD, which has been previously shown in studies involving living subjects.^19^ In the CSF-/PET+ discordant case we presented, the patient with a clinically advanced AD dementia had a CSF Aβ_42_ value just above the cut-off, but CSF p-tau/Aβ_42_ ratio was in the pathological range. In this case, CSF Aβ_42_ was likely false-negative, possibly due to individual differences in CSF dynamics, as both CSF t-tau and p-tau were already increased. This also highlights the advantage of using continuous measurements as opposed to binarized data, as the distance from cut-off includes additional information. Although (p)tau/Aβ_42_ ratio may be superior to Aβ_42_ when predicting clinically advanced disease with increased (p)tau levels, this may hamper the detection of merely amyloid-β pathology, where tau tangle pathology has not yet begun. This may become clinically significant if anti-amyloid treatment arrives in the future. Finally, we reported an isolated increase of CSF t-tau with normal CSF ptau levels in three non-AD cases (two FTLD, one HDLS). Although CSF t-tau and p-tau are highly correlated, this finding supports the notion that CSF t-tau can increase in other brain pathologies^46^ and CSF p-tau is more AD-specific.^47^

The primary strength of our study is the availability of two amyloid-β biomarkers and a neuropathological assessment in a relatively large patient cohort that allowed us to compare the two *in vivo* amyloid-β biomarkers to neuropathological change. Although PET and CSF were usually performed close in time, there was a median 3-year delay between the amyloid-β biomarkers and autopsy, as is often the case with studies involving *in-vivo* biomarkers and autopsy data. While this might have impacted our results, a major change over three years is unlikely, given the remarkably slow course of AD.^48^ We used standardized uptake value ratio images for PET visual read, which could have an impact on our results as non-displaceable binding potential images have been shown to be more reliable in detecting early amyloid-β pathology.^49,50^ Another limitation is that we included subjects from the year 2006, and over time technologic advancement has taken place, leading to both increased image quality of PET scans and understanding of pre-analytical factors influencing CSF (leading to longitudinal drift of median values, in our cohort).

In conclusion, our findings illustrate a range of reasons for the amyloid-β CSF/PET discordance, and that even concordant amyloid-β biomarker positivity accurately reflecting amyloid-β pathology does not always equal a definite neuropathological diagnosis of AD. Thus, it is important to consider co-morbidities as well as other neurodegenerative diseases when using amyloid-β biomarkers for clinical diagnosis, especially since molecular biomarkers for non-AD neurodegenerative diseases are currently lacking.

## Data Availability

Data is not publicly available.

## Acknowledgements

The Alzheimer Center Amsterdam is supported by Alzheimer Nederland and Stichting VUmc fonds. Research performed at the Alzheimer Center Amsterdam is part of the neurodegeneration research program of Amsterdam Neuroscience. JR would like to thank Sergei Nazarenko, the International Atomic Energy Agency (IAEA) and the North Estonia Medical Centre for their contribution to his professional development.

## Authors’ contributions

JR, RO, FB and PS conceived the study and designed the protocol. JR performed statistical analysis, analyzed/interpreted data and drafted the manuscript. RO and FB provided overall study supervision. JR, BB, LC, RO, FB participated in writing the manuscript. LC and BvB did the visual reads of the amyloid-β PET scans. BB and AR were responsible for neuropathological evaluation. CT, AR, BvB, PS had a major role in the acquisition of data, and critically revised and edited the manuscript for intellectual content. All authors read and approved the final version of the manuscript.

